# Non-Invasive Prenatal Testing (NIPT) Results for Participants of the eXtraordinarY Babies Study: Screening, Counseling, Diagnosis, and Discordance

**DOI:** 10.1101/2021.11.03.21265829

**Authors:** Susan Howell, Shanlee M Davis, Talia Thompson, Mariah Brown, Tanea Tanda, Karen Kowal, Amanda Alston, Judith Ross, Nicole R Tartaglia

## Abstract

Sex chromosome aneuploidies (SCAs), including 47,XXY, 47,XXX, 47,XYY, and supernumerary variants, occur collectively in approximately 1/500 livebirths. Clinical phenotypes are highly variable resulting in previous ascertainment rates have been estimated to be only 10-25% during a lifetime. Historically, prenatal SCA diagnoses were incidental findings, accounting for ≤10% of cases, with the majority of diagnoses occurring postnatally during evaluations for neurodevelopmental, medical, or infertility concerns. The initiation of noninvasive prenatal testing (NIPT) in 2012 and adoption into standardized obstetric care provides a unique opportunity to significantly increase prenatal ascertainment of SCAs. However, the impact NIPT has had on ascertainment of SCAs is understudied, particularly for those who may defer diagnostic testing until after birth. This study evaluates the timing of diagnostic testing following positive NIPT in 152 infants with SCAs and potential factors influencing this decision. Eighty-seven (57%) elected to defer diagnostic testing after a positive NIPT until birth, and 8% (7/87) of those confirmed after birth were found to have discordant results on postnatal diagnostic testing, most of which would have influenced genetic counseling.

## INTRODUCTION

Sex chromosome aneuploidies (SCAs), including Klinefelter syndrome/47,XXY, Trisomy X/47,XXX, 47,XYY syndrome, and 48,XXYY syndrome, are the most prevalent supernumerary chromosomal conditions, occurring collectively in approximately 1/500 livebirths. Clinical phenotypes are highly variable in these conditions, often with mild dysmorphic features or neurodevelopmental involvement, resulting in only 10-25% lifetime ascertainment (Abramsky & Chapple, 1997). Historically, prenatal SCA diagnoses accounted for 10% or less of SCA cases and were often incidental findings following CVS or amniocentesis for advanced maternal age, and the majority of SCA diagnoses occurred in the postnatal period during clinical evaluations for neurodevelopmental, medical, or infertility concerns (Bojesen, Juul, & Gravholt, 2003). The initiation of cell-free fetal DNA screening, commonly referred to as noninvasive prenatal testing (NIPT), in 2012 and subsequent adoption into standardized obstetric care, has drastically changed the landscape for prenatal identification of chromosomal abnormalities. This has provided a unique opportunity to identify SCAs prenatally (Wilson et al., 2013). Beginning in 2016, and most recently updated in 2020, the American College of Obstetrics and Gynecology issued a position statement recommending that NIPT be universally offered to all pregnant women, regardless of a priori risk, as it is a superior screening test to other alternatives citing the highest level of evidence (Gregg et al., 2016; “Screening for Fetal Chromosomal Abnormalities: ACOG Practice Bulletin Summary, Number 226,” 2020). These guidelines also state that all patients with a positive NIPT should receive genetic counseling and be offered diagnostic testing via chorionic villus sampling or amniocentesis to confirm these screening results.

With the utilization and growing adoption of NIPT, prenatal ascertainment rates of SCAs and subsequent number of infants known to have SCAs, are logically anticipated to rise. This opportunity led to the development of the eXtraordinarY Babies Study, a prospective natural history study of infants prenatally identified and subsequently diagnosed with SCA designed to examine trajectories of neurodevelopment and physical health from birth through the first few years of life as well as psychosocial factors including quality of life and parental experiences. Funded by the National Institute of Child Health and Human Development (NICHD) and in collaboration with the American College of Medical Genetics and Genomics (ACMG) Newborn Screening Translational Research Network (NBSTRN) (ClinicalTrials.gov NCT03396562), the eXtraordinarY Babies Study enrolls infants between 2 and 12 months of age with a prenatal result (NIPT or diagnostic) of SCA, with longitudinal evaluations conducted at two sites including University of Colorado/Children’s Hospital Colorado and Nemours-Dupont Hospital for Children. While the eXtraordinarY Babies Study aims to prospectively describe and compare the natural history of SCA conditions, identify predictors of outcomes in SCA, and build a rich data set linked to a biobank for future study, much has also been learned about diagnostic testing outcomes following NIPT results positive for SCA.

Historically, most studies evaluating outcomes following NIPT often limit follow up to the gestational period. One report found that NIPT has not increased the prevalence of infants known to have SCAs at birth, although this study only included cases with confirmed prenatal diagnostic genetic testing (Howard-Bath, Poulton, Halliday, & Hui, 2018). Given maternal pregnancy history, procedural risks inherent in prenatal diagnostic testing and other factors, women may elect to defer diagnostic testing until after birth. As such, studies evaluating the overall impact NIPT has made to increasing ascertainment of SCAs need to include both pre- and postnatal diagnostic testing results following an NIPT result positive for SCA.

Prenatal genetic counseling for SCA-positive NIPT results is challenged by relatively poor positive predictive values for SCAs in NIPT, highly variable phenotypic outcomes, and historic peer-reviewed publications inherently biased by ascertainment (Mennuti, Chandrasekaran, Khalek, & Dugoff, 2015; Petersen et al., 2017; Wang et al., 2020). While NIPT has been demonstrated to have high sensitivity and specificity in identification of other chromosomal conditions, such as Trisomy 21/Down syndrome, the positive predictive values (PPV) for the detection of SCAs have varied from 25-89% and many companies fail to include these test statistics for SCAs on their result reports entirely (Lu et al., 2021; Shi et al., 2021; Skotko et al., 2019; Zheng et al., 2020). Phenotypes among SCAs range widely from mild dysmorphisms and tall stature to increased rates of cognitive impairment, medical conditions and psychological features. Furthermore, genetic counseling for SCAs is reliant upon peer-reviewed literature publications, the majority of which include data from individuals who were postnatally ascertained due to presenting neurodevelopmental, medical or fertility problems. As such, parental decision making for pursuing prenatal diagnostic testing at the time of an NIPT result may be overshadowed by anxiety and psychological distress balanced by decisional conflict, especially in consideration of inherent prenatal diagnostic procedural risks (Labonte, Alsaid, Lang, & Meerpohl, 2019; Lewis, Hill, & Chitty, 2016). In one retrospective study of 61 cases with positive NIPT for trisomy SCAs, only 24% elected to have prenatal diagnostic testing (Ramdaney, Hoskovec, Harkenrider, Soto, & Murphy, 2018). Factors affecting the decision for timing of diagnostic testing rely upon the personal history of the mother as well as information provided at the time of the result. The professional providing information and whether the identified condition was discussed prior to testing may also influence this decision (Fleddermann et al., 2019; Marteau et al., 2002; Riggan, Close, & Allyse, 2020; Sadlecki, Grabiec, Walentowicz, & Walentowicz-Sadlecka, 2018). This is especially important to consider for the SCA conditions, as most SCAs are often not discussed during pretest consent and even more surprising due to lower public knowledge of SCT compared to Down syndrome.

Counseling for NIPT results positive for SCA are typically directed to the condition reported, yet given the complexities of interpretation in SCA NIPT, discordant abnormal diagnostic results should be considered in counseling as well (Ramdaney et al., 2018). This paper aims to report on 152 participants from the eXtraordinarY Babies Study with SCA initially identified by NIPT, the parental decisions for diagnostic testing, and parent perceptions of providers’ knowledge and quantity of information presented following a positive NIPT result. We also report a series of abnormal discordant diagnostic outcomes to further inform prenatal genetic counseling for NIPT results positive for SCAs.

## METHODS

Participants of this study provided informed written consent for the eXtraordinarY Babies Study (Colorado COMIRB#17-0118; Nemours Office of Human Subjects Protection #1151006; NIH/NICHD# R01HD42974; ClinicalTrials.gov# NCT03396562). Inclusion in the eXtraordinarY Babies Study requires prenatal identification of an SCA either by NIPT or diagnostic prenatal testing, with confirmatory cytogenetic testing conducted prenatally and/or postnatally if NIPT, and enrollment between 6 weeks and 13 months of age. Exclusion criteria include birth <34 weeks, presence of an additional genetic or metabolic disorder with neurodevelopmental or endocrine involvement, presence of a congenital malformation (not previously described with SCA), or neonatal complications such as hypoxic-ischemic brain injury or neonatal meningitis. This analysis includes participants of the eXtraordinarY Babies Study who were prenatally identified by NIPT with subsequent diagnostic cytogenetic testing (prenatal and/or postnatal) and who had provided reports from both tests for review. Participants were excluded from this analysis if either NIPT reports or diagnostic test results could not be obtained, or if their prenatal diagnosis was first identified by amniocentesis or CVS. A total of 152/255 total participants enrolled in the eXtraordinarY Babies Study were included in this analysis. Data abstracted from the eXtraordinarY Babies Study included demographic information (socioeconomic, race, ethnicity, state of residence to identify geographic region), family history (maternal date of birth to calculate age at delivery, maternal height and maternal pre-pregnancy weight to calculate pre-pregnancy BMI) and birth history (date of birth, gestational age, birth weight). NIPT reports were reviewed and abstracted by a board certified genetic counselor (SH) to record commercial lab, date of sample collection (which was then used to calculate gestational age at the time of sample collection based on gestational age at the date of birth), date of NIPT result report, fetal fraction, and sensitivity, specificity, and positive predictive value for SCA (if reported). A one-page questionnaire was completed by 102/152 parents of participants, self-reporting date and child’s age at the time of questionnaire completion and the following additional information:

– Gestational age at the time of SCA identified/diagnosed
– Type(s) of prenatal testing which identified the diagnosis
– Reason(s) prenatal testing was performed
– Medical provider’s specialty who ordered prenatal screening/testing
– If the mother was informed about possible SCA diagnosis at the time of NIPT consent
– What provider(s) informed mother about the SCA diagnosis
– If the mother met with a genetic counselor after receiving results (NIPT and/or prenatal diagnostic testing)
– How and what type of information about the SCA was provided
– The perceived amount of information provided
– If the provider was perceived to be well-informed about the SCA condition
– If the diagnosis was confirmed after birth and if so, were the results the same as prenatally identified.

Analysis of these results were conducted by descriptive statistics and SPSS utilizing chi-square analysis or r-value (for continuous variables) to calculate *p* values for significance (p<0.05).

## RESULTS

Demographics for the 152 infants analyzed in this cohort (104 XXY, 27 XXX, 15 XYY, and 6 XXYY) are shown in Table 1. Over half (57%) delayed diagnostic testing until after birth, of which 85% (postnatally confirmed) occurred prior to 2 months of age. We found no difference between timing of diagnostic testing based on maternal age, race, ethnicity, geographic region, self-reported indications for pursuing NIPT, maternal health history, family history, abnormal ultrasound findings, SCA karyotype result, or PPV for NIPT results. Participants earning less than $100k were less likely to pursue prenatal confirmatory testing than those in higher income brackets (>$250k, p=.010; $150-$250k, p=.017), although differences were not statistically significant after adjustments were made for multiple comparisons. Of the 43% (n=65/152) of participants who pursued prenatal diagnostic testing following NIPT, 80% elected an amniocentesis procedure. Details of elected procedures, timing of diagnostic testing, comparisons of characteristics between those deferring to postnatal testing, and prenatal counseling experience questionnaire results are presented in Tables 2 and 3.

**Table 1:** Subject Demographics.

| Table 1: Subject Demographics |  |
| --- | --- |
|  | Total (N= 152) |
| **Race** |  |
| Caucasian | 138 (91%) |
| Asian | 11 (7%) |
| Native American | 3 (2%) |
| African American | 2 (1%) |
| **Ethnicity** |  |
| Non-Hispanic | 134 (88%) |
| Hispanic | 18 (12%) |
| **Annual Household Income** |  |
| <\$100k | 42 (29%) |
| \$100k-\$150k | 36 (25%) |
| \$150k-\$250k | 39 (27%) |
| >\$250k | 23 (16%) |
| **Gestational age at NIPT (wks)** | 13 ± 5.4 |
| **Fetal Fraction on NIPT (%)** | 9.2 ± 3.9 |
| **Maternal pre-pregnancy BMI** | 26 ± 6.0 |
| **Reason reported for NIPT** |  |
| Maternal Age | 59 (57.8%) |
| Abnormal Ultrasound | 5 (4.9%) |
| Routine / Elective / Learn Gender | 43 (42.1%) |
| Other (family history, prior pregnancy loss) | 10 (9.8%) |
| **Prenatal Diagnostic testing (CVS or Amniocentesis)** | 65 (43%) |
| **Final Karyotype Result** |  |
| 47,XXY | 104 (68%) |
| 47,XXX | 27 (18%) |
| 47,XYY | 15 (10%) |
| 48,XXYY | 6 (4%) |
| **Birthweight (kg)** | 3.25 ± 0.7 |
| **Gestational age at Delivery (wks)** | 38.7 ± 1.4 |
| **Maternal Age at Delivery (yrs)** | 35.3 ± 4.8 |
| **Geographic Region<sup>†</sup>** |  |
| Northeast | 31 (20%) |
| Midwest | 24 (16%) |
| South | 42 (28%) |
| West | 53 (35%) |
| International | 2 (1%) |

<sup>†</sup>US Geographic Regions as designated by the US Census Bureau: Northeast: CT, MA, ME, NH, NJ, NY, PA, RI, VT; Midwest: IA, IL, IN, KS, MI, MN, MO, NE, ND, OH, SD, WI; South: AL, AR, DC, DE, FL, GA, KY, LA, MD, MS, NC, OK, SC, TN, TX, VA, WV; West: AK, AZ, CA, CO, HI, ID, MT, NM, NV, OR, UT, WA WY
Note: Data are mean ± standard deviation (SD) or n (%)

**Table 2:** Karyotypes Stratified by Timing of Diagnostic Testing.

| **Table 2: Karyotypes Stratified by Timing of Diagnostic Testing** |  |  |  |  |
| --- | --- | --- | --- | --- |
| **Diagnostic Karyotype** | **Full cohort of NIPT Positive for SCA† (n=152)** | **CVS Prenatal Diagnostic Testing (n=13)** | **Amnio Prenatal Diagnostic Testing (n=52)** | **Deferred to Postnatal Diagnostic Testing (n=87) ‡** |
| **47,XXY** | 104 (68%) | 10 (10%) | 37 (36%) | 57 (55%)† |
| **47,XXX** | 27 (18%) | 1 (4%) | 11 (41%)† | 15 (56%) |
| **47,XYY** | 15 (10%) | 2 (13%) | 3 (20%) | 10 (67%) |
| **48,XXYY** | 6 (4%) | - | 1 (17%)† | 5 (83%) † |
| **All SCAs** | 152 (100%) | 13 (9%) | 52 (34%) | 87 (57%) |

† Includes Discordant NIPT vs. Diagnostic Results, see Table 4
‡ 85% of all diagnostic confirmations deferred to after birth were conducted prior to 2 months of age and were confirmed in cord blood or peripheral blood

**Table 3:**
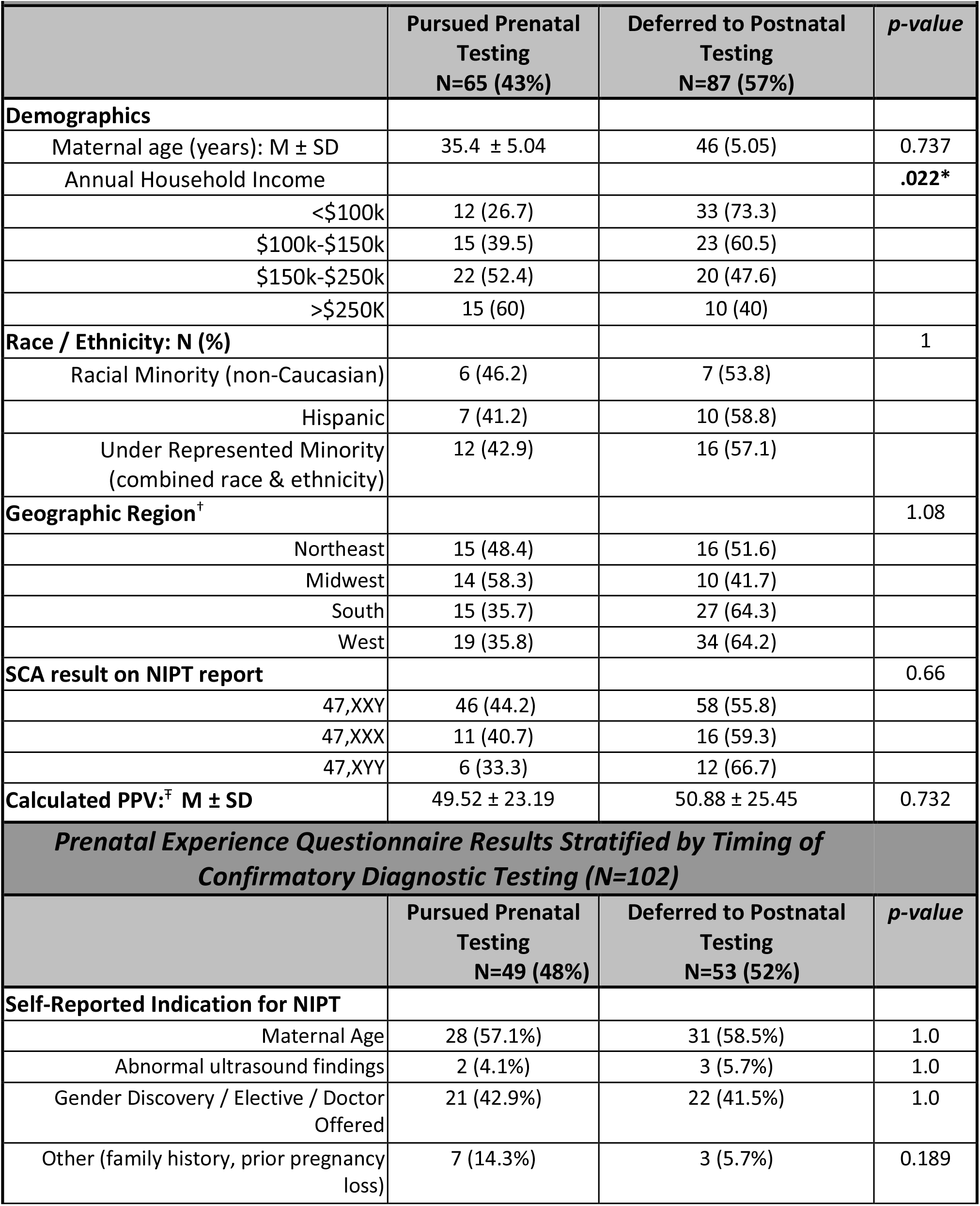
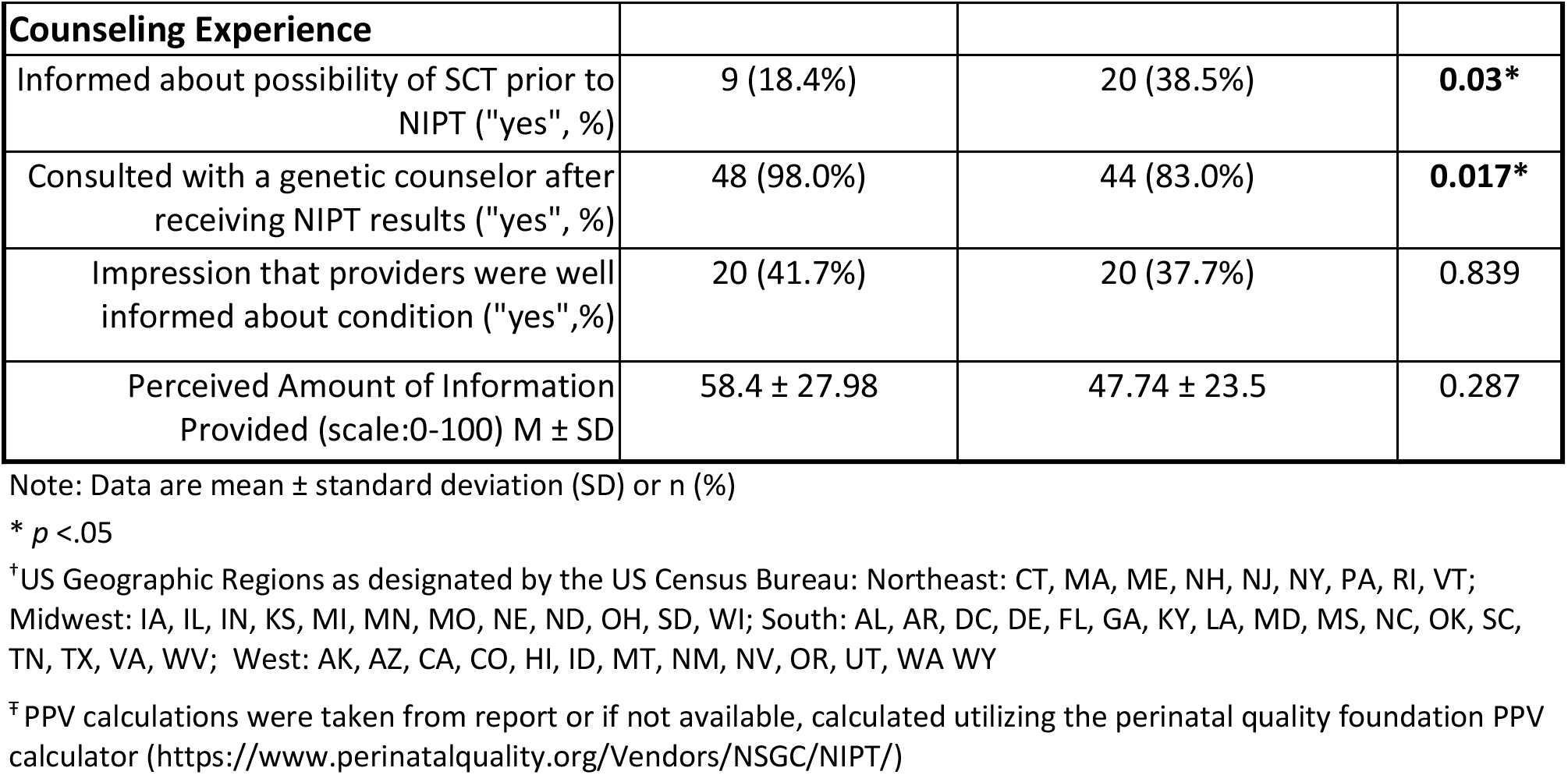
Participant Characteristics Stratified by Timing of Confirmatory Diagnostic Testing.

Eleven (7%) diagnostic results were discordant with NIPT results. Of these, 2 participants were found to be mosaic with a typical cell line, while 9 participants had a different SCA condition altogether. Seven of these 9 participants with discordant results had deferred diagnostic testing until birth. Details regarding fetal fraction on NIPT, maternal age at delivery, maternal pre-pregnancy BMI, and diagnostic test pursued for these 11 participants with NIPT results discordant from diagnostic results are presented in Table 4.

**Table 4:**
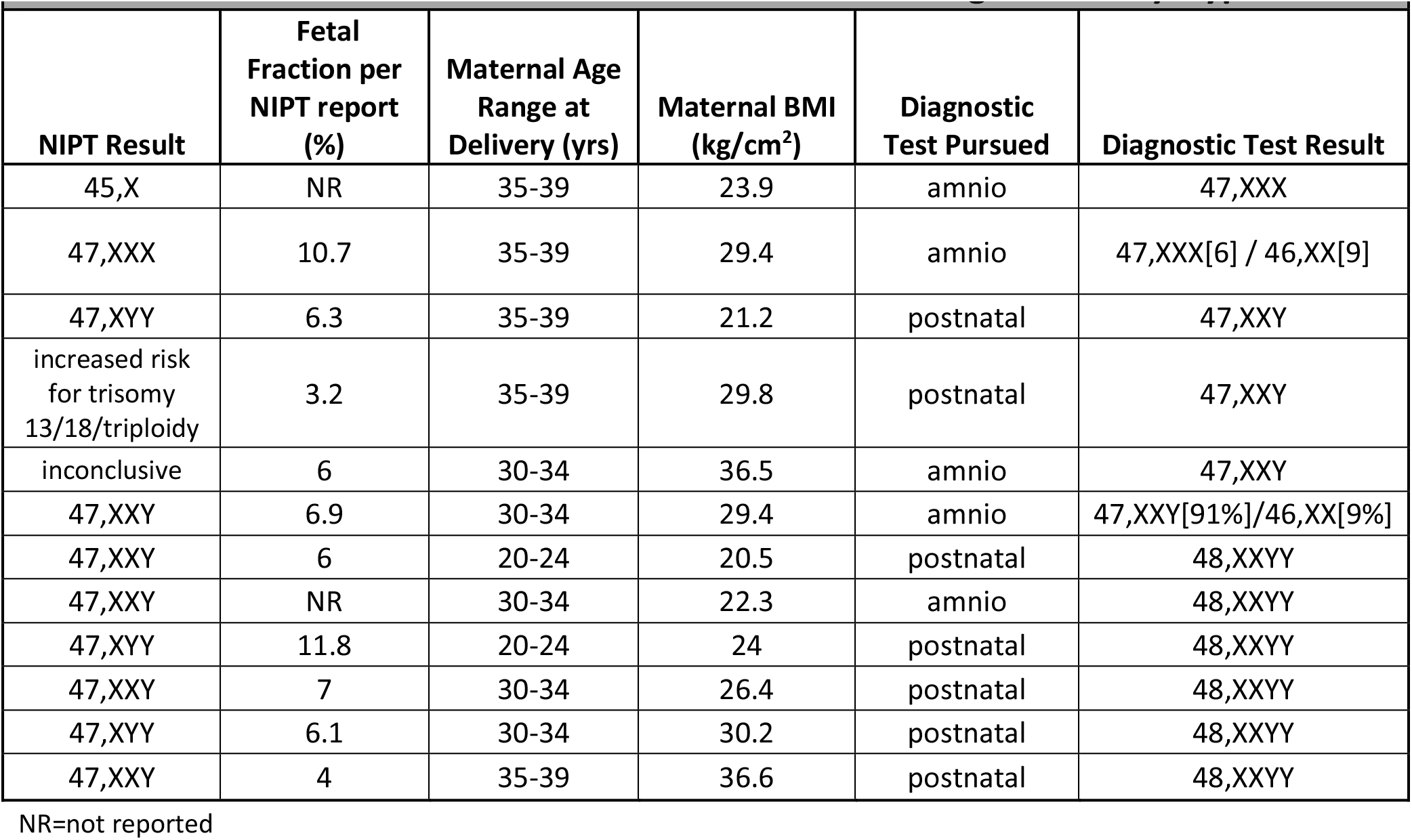
Discordant Results Between NIPT and Diagnostic Karyotype.

Of the 152 total participants included for this study, 102 participants completed a one-page questionnaire self-reporting reasons for NIPT, experiences with prenatal genetic counseling and potential counseling factors influencing diagnostic testing decisions (see Table 3). The top two indications reported for pursuing NIPT were maternal age (58%) and elective/gender discovery/doctor offered (48%). The majority of participants consulted with a genetic counselor (90%) after receiving their results (NIPT and/or prenatal diagnostic test results) and those who pursued prenatal diagnostic testing were more likely to have received genetic counseling (p=.017) Participants who were informed of the possibility of SCA prior to NIPT were significantly more likely to defer to postnatal diagnostic testing (p=.03). While we found no difference in diagnostic timing based on perceptions of the amount of information provided or how well-informed providers counseling were about the SCA, less than half of participants felt their provider(s) were “well-informed” about the SCA discussed and participants who endorsed their provider was “well-informed” reported receiving significantly more information than those who endorsed their provider was not well-informed (p<.001).

## DISCUSSION

The majority of studies on NIPT results positive for SCA focus on the analytical performance of the test limited to prenatal outcomes. In this study, we present 152 cases of NIPT results positive for SCA with their diagnostic testing results, identifying over half of these parents delayed diagnostic testing until after birth. However, 7% (11/152) of NIPT results positive for SCA were discordant with the diagnostic test result, with 9 of these 11 results warranting different genetic counseling than what would be indicated based on the NIPT results alone (i.e trisomy SCA versus tetrasomy SCA). Further, 7 of these 9 discordant results elected to defer to postnatal diagnostic testing, likely based on counseling provided in conjunction with additional fetal anatomy ultrasound (Fleddermann et al., 2019). However, sex chromosome trisomies are infrequently associated with second trimester ultrasound findings, so it is unlikely that ultrasound markers to modify the PPV will be recognized (De Vigan et al., 2001). This study highlights that NIPT results positive for SCA are often deferred for diagnostic testing postnatally, that families benefit from receiving more information which results in feeling that the provider counseling is well-informed about the SCA condition being discussed, and that counseling for NIPT results should address the possibility of discordance among NIPT and diagnostic SCA potential results.

A 2018 international population-based study concluded that while SCAs contribute to a higher percentage of confirmed prenatal diagnoses secondary to NIPT, the decline in prenatal diagnostic testing leads to a relatively steady prevalence of prenatally confirmed SCAs (Howard-Bath et al., 2018). The findings of our study demonstrate that less than 50% of pregnancies with NIPT results positive for SCA pursue prenatal diagnostic testing but the majority rather defer diagnostic testing to the postnatal period. Other studies have shown even lower percentages (25-34%) of mothers who pursue prenatal diagnostic testing after an NIPT result positive for SCA (Ramdaney et al., 2018; Riggan et al., 2020). These high rates of deferral to postnatal diagnostic testing emphasize that estimates of the impact from NIPT on the ascertainment of SCAs should include both prenatal and postnatal diagnostic testing. As the study by Howard-Bath et al in 2018 demonstrated a steady birth prevalence of SCAs based on prenatal diagnostic testing after NIPT, based on our results and similar studies suggesting 60-80% of those receiving positive NIPT results will have diagnostic testing shortly after birth, it can be estimated that introduction of NIPT has increased the overall SCA ascertainment in infancy by at least two to three-fold. Anecdotally, we appreciate this in our clinical practice, however additional population-based studies are needed to confirm these assumptions.

While we collected a limited data set of potential factors influencing the decision to defer to postnatal diagnostic testing, our study did identify a significant difference in deferral to postnatal confirmation when SCA was discussed prior to NIPT. This finding could be attributed to implicit framing effects during pre-NIPT genetic counseling, especially in context of counseling for all possible NIPT outcomes, which could precipitate post-NIPT decision making (van der Steen et al., 2019). In addition, we also found a difference approaching significance in deferral to postnatal testing when mothers reported an annual household income less than $100k, which warrants further investigation. While our study did not inquire as to how or why socioeconomic factors influenced diagnostic decision making, previous research has demonstrated socioeconomic disparities in prenatal genetic screening and informed decision making due to limited access to care or information provided during counseling (Khoshnood, Blondel, de Vigan, & Breart, 2004). We did not find any differences in our results based on race or ethnicity, however our study sample size is limited and further investigation is warranted as previous research evaluating racial/ethnic groups with NIPT results positive for SCAs identified that African American women were the most likely to decline prenatal diagnostic testing, while Asian women were the most likely to elect for prenatal diagnostic testing (Ramdaney et al., 2018). These collective findings and insights highlight the need for future research further investigating disparities in prenatal genetic counseling and testing for SCAs, possible reasons for these disparities, and how to minimize them.

A unique aspect of prenatal genetic counseling following an NIPT result positive for SCA is the presentation, interpretation and often calculation of the positive predictive value (PPV). The PPV for NIPT results regarding SCAs is inherently variable among laboratories with published values ranging from 20-86% (Lu et al., 2021; Petersen et al., 2017; Ramdaney et al., 2018; Shi et al., 2021). A 2019 review of 10 NIPT laboratory reporting methods concluded recommendations that laboratory reports visibly and clearly state the detection rate (DR), specificity (SPEC), positive predictive value (PPV), and negative predictive value (NPV) for all conditions being screened in order to assist patients and providers in making decisions and interpreting results (Skotko et al., 2019). As noted in the review, no commercial labs published their PPV on the respective reports (at that time). While some improvements have been made to various lab reports since this 2019 publication, there continues to be significant variability in what information is disclosed on NIPT result reports industrywide and many omit PPV values. Recognizing the importance of these variables for clinical interpretation and informed counseling, a taskforce was established, including members of the National Society of Genetic Counselors and the Perinatal Quality Foundation, to review the medical literature and build consensus regarding best estimates to develop algorithms and ultimately the publication of the NIPT/cffDNA Predictive Value Calculator (https://www.perinatalquality.org/Vendors/NSGC/NIPT/). When estimates of sensitivity and specificity are not provided on the lab report, this calculator utilizes estimates based on a meta-analysis of available studies (Gil, Quezada, Revello, Akolekar, & Nicolaides, 2015). Today, the NIPT/cffDNA Predictive Value Calculator published by the PerinatalQuality Foundation and the National Society of Genetic Counselors (NSGC; www.perinatalquality.org) provides genetic counselors with a tool to estimate the PPV when faced with an NIPT result positive for SCA. While this calculator is intended to facilitate informed decision making, counseling for SCAs commonly results in setting expectations of a false positive if PPV is below 50%, in which mothers perceive diagnostic lab outcomes more likely to be normal. While we did not find any difference in timing of diagnostic testing based on PPV (provided by the lab or calculated online), further studies evaluating mothers’ expectations based on presented PPV may be useful to improve genetic counseling when NIPT results are positive for SCA given the relatively poor PPVs.

While consent for NIPT testing may be influenced by various factors ranging from desire of early fetal gender identification to experiences of previous pregnancy outcomes, NIPT results positive for SCA may have a pivotal psychological impact on the expectant mother. In a 2013 study by Lalatta et al., the importance of utilizing a framework in genetic counseling, including the potential findings for SCAs, prior to prenatal diagnosis was supported to help reduce the emotional devastation with unexpected results of SCA given the relatively high incidence of these conditions compared to other aneuploidies (Lalatta & Tint, 2013; Riggan et al., 2020). While women who receive an NIPT result positive for SCA are recommended to receive genetic counseling regarding diagnostic testing options, the approach to prenatal genetic counseling for SCAs still continues to be far from standardized (Gregg et al., 2016). In a 2019 study surveying 176 genetic counselors to evaluate genetic counseling practices throughout the United States following an NIPT result positive for SCA, significant discrepancies were identified that highlighted the need to establish professional guidelines in order to provide consistencies in care for NIPT results positive for SCA (Fleddermann et al., 2019).

Effective prenatal genetic counseling is fundamental in providing accurate, unbiased, and updated information alongside nondirective psychological support for families faced with an at-risk or confirmed prenatal genetic diagnosis. As such, it is imperative to evaluate the prenatal genetic counseling experiences and diagnostic timing decisions in parents who continued pregnancies following NIPT results positive for SCAs. The majority of participants in our study reported that they met with a genetic counselor after receiving results (NIPT or diagnostic),yet less than 50% of participants felt their provider was “well-informed” about SCAs. We found no difference in decisions in timing of diagnostic testing based on the amount of information provided about the SCA. Our study results did demonstrate that consultation with a genetic counselor after results were received was associated higher likelihood of prenatal diagnostic testing, and the amount of information provided during genetic counseling was positively and significantly associated with mothers’ perceptions that providers were well-informed (r_pb_ = .598, n=101, p<.001). These findings are consistent with previous publications reporting that even genetic providers feel poorly equipped to provide adequate support at the time of SCA counseling based on limited time during appointments, lack of knowledge regarding SCAs and few educational resources available (Farrell, Agatisa, Mercer, Mitchum, & Coleridge, 2016; Riggan et al., 2020). Our study promotes future comprehensive education programs regarding SCA for genetic counselors and the importance of extensive information regarding SCA be provided to mothers at the time of counseling in order to appropriately support informed decision making.

Importantly, our study presents a series of 11 participants (7% of our total sample) in which NIPT SCA results were discordant with the final SCA diagnosis, of which 9 participants were diagnosed with a different condition and would have been counseled inaccurately if counseled based upon the NIPT result condition alone. Discordance between NIPT result and fetal karyotype has been well established to be attributed by various factors including, but not limited to, confined placental mosaicism, maternal copy number variation (CNVs), maternal X chromosome aneuploidy and/or mosaicsm, maternal malignancy, vanishing twin, and technical, bioinformatics, or human errors (Hartwig, Ambye, Sorensen, & Jorgensen, 2017). For these and other reasons, NIPT remains classified as a screening, non-diagnostic test with standard recommendations that any positive NIPT result be followed by confirmatory diagnostic testing (Devers et al., 2013; Hartwig et al., 2017). However, five of our 9 discordant results showed an NIPT result for a sex chromosome trisomy (XXY or XYY) and parents elected to defer to postnatal diagnostic testing, which subsequently resulted in an unexpected diagnosis of a tetrasomy, 48,XXYY. Similarly, a retrospective study of 27 NIPT screens positive for XXY had discordant results with other SCAs (XYY, XXYY, and XXXXY) upon diagnostic testing (Ramdaney et al., 2018). While postnatal recall of prenatal counseling experiences has inherent limitations and biases, routine counseling for NIPT results of XXY or XYY does not routinely provide in-depth information regarding a possible diagnosis of 48,XXYY (or other tetrasomy outcomes to facilitate informed decision making. Traditionally, genetic counseling for NIPT results is based upon the presenting NIPT lab report. These 5 discordant results represent the imperative need for prenatal genetic counseling on NIPT results positive for SCAs to also include the possibility for an SCA diagnosis that is abnormal but discordant with the NIPT lab result. In a recent study this concern is articulated specific to NIPT results positive for 47,XXY, with the authors underlining the importance of a definitive diagnosis not only for excluding a false positive but also excluding other chromosomal variations which may have a different and more severe phenotype (Ronzoni et al., 2020). Our study findings reinforce the importance of counseling regarding possible other SCAs as there are significant phenotypic differences associated with higher risks of medical complexity and neurodevelopmental involvement when comparing sex chromosome trisomies vs. pentasomies, such as 48,XXYY (Raznahan et al., 2018; Skuse, Printzlau, & Wolstencroft, 2018; N. Tartaglia, Ayari, Howell, D’Epagnier, & Zeitler, 2011; N. R. Tartaglia, Ayari, Hutaff-Lee, & Boada, 2012). By providing comprehensive counseling regarding interpretation of NIPT results and possible outcomes, couples are able to make a more-informed decision in context of additional factors that impact the PPV in NIPT results positive for SCA including the potential for a discordant SCA diagnosis and associated outcomes.

Limitations in this study included only pregnancies which were continued after NIPT screening and confirmed to have an SCA condition, timing of postnatal testing may have been influenced by the desire to meet inclusion criteria of the eXtraordinarY Babies Study (confirmatory diagnosis of SCA prior to 13 months of age), and intentional or unintentional prenatal recall bias on postnatal questionnaires.

In consideration of future areas of research, investigation of possible reasons for disparities in prenatal genetic testing in SCA and how to minimize these disparities is warranted. Additionally, studies are needed to better inform genetic counselors about SCA and potential discordant outcomes when NIPT results are positive. Recognizing the phenomenon of some mothers pursuing prenatal diagnostic testing, while other mothers defer testing to after birth, results in a two-tier ascertainment impact from NIPT screening in SCA. Future areas of research could further investigate if the postnatal outcomes in the children or if the parental experiences, such as attachment, differ significantly among these two cohorts. Additional areas for future research could include investigation into the long-term emotional health of parents raising a child with an SCA initially identified by NIPT, including discordant results, and prenatal genetic counseling factors that impacted these parental outcomes.

In conclusion, our study supports that the majority of NIPT results positive for SCA are confirmed postnatally, that NIPT has increased the ascertainment of SCAs two-to three-fold when accounting for both prenatal and postnatal diagnostic tests, and that prenatal counseling for NIPT results positive for SCA should include providing extensive information regarding the SCA and discussion regarding possible abnormal but discordant diagnostic outcomes in order for mothers to feel well-informed and able to make an informed decision regarding diagnostic testing.

## Data Availability

Deidentified data that support the findings of this study are available from the corresponding author upon reasonable request.

## ACKNOWLEDGEMENTS

The authors wish to thank the study participants in the eXtraordinarY Babies Study and their families. This work was also supported by NIH/NCATS Colorado CTSA Grant Number UL1 TR002535, NIH/NINDS K23NS070337, NIH/NICHD K23HD092588, and NIH 2RO1-HD42974. Contents are the authors’ sole responsibility and do not necessarily represent official NIH views.

